# Active Neural Networks to Detect Mentions of Changes to Medication Treatment in Social Media

**DOI:** 10.1101/2020.12.04.20244210

**Authors:** Davy Weissenbacher, Suyu Ge, Ari Klein, Karen O’Connor, Robert Gross, Sean Hennessy, Graciela Gonzalez-Hernandez

## Abstract

**Objective:** We address a first step towards using social media data to supplement current efforts in monitoring population-level medication non-adherence: detecting changes to medication treatment. Medication treatment changes, like changes to dosage or to frequency of intake, that are not overseen by a physician are, by that, non-adherence to medication. Despite the consequences, including worsening health conditions or death, 50% of patients are estimated to not take medications as indicated. Current methods to identify non-adherence have major limitations. Direct observation may be intrusive or expensive, and indirect observation through patient surveys relies heavily on patients’ memory and candor. Using social media data in these studies may address these limitations.

**Methods:** We annotated 9,835 tweets mentioning medications and trained a convolutional neural network (CNN) to find mentions of medication treatment changes, regardless of whether the change was recommended by a physician. We used active and transfer learning from 12,972 reviews we annotated from WebMD to address the class imbalance of our Twitter corpus. To validate our CNN and explore future directions, we annotated 1,956 positive tweets as to whether they reflect non-adherence and categorized the reasons given.

**Results:** Our CNN achieved state-of-the-art performance with 0.50 F1-score. The manual analysis of positive tweets revealed that non-adherence is evident in a subset with nine categories of reasons for non-adherence.

**Conclusion:** We showed that social media users publicly discuss medication treatment changes and may explain their reasons including when it constitutes non-adherence. This approach may be useful to supplement current efforts in adherence monitoring.

## 1. Introduction

Medication non-adherence refers to when patients do not follow medication treatments as prescribed by their doctors. Non-adherence can be subdivided into three categories (De Geest, et al., 2018). In primary non-adherence, patients do not fill their prescriptions or do not start their treatments. In non-persistence, patients stop their treatments, intentionally or unintentionally, without being advised by a health professional to do so. In suboptimal execution, patients are taking their medications but not as recommended (ex. wrong dosage or frequency).

Medication non-adherence has long been recognized as a major contributor to health problems (Reddy, 2012; Vrijens, et al., 2012), with the first mention dating from Hippocrates (Reddy, 2012; Vrijens, et al., 2012; Hippocrates, 1923). In 2003, the WHO estimated than 50% of patients in developed countries were failing to follow their medical treatment. In 2018, non-adherence led to an estimated 275,689 deaths at an annual cost of $528 billion per year in the United States alone (Watanabe, et al., 2018). Understanding its causes may help us design effective interventions to improve adherence (Hugtenburg, et al., 2013; Marcum, et al., 2013). According to Osterberg and Blaschke (Osterberg & Blaschke, 2005), the real barriers to adherence lie in deficient interactions between patients, providers, and the health care system. For example, by recommending a complex treatment, a provider increases the probability that the patient would skip a medication; by maintaining high cost for a medication, a health care system increases the risk for the patient to not refill a prescription; and by showing poor knowledge of medication costs, a provider may prescribe expensive drugs even when more affordable alternatives are available. Furthermore, in clinical practice, patients rarely reveal their non-adherence. And even if they do, they may be reluctant to openly discuss with their healthcare providers the true reasons for altering their therapy against the providers’ advice (Yin Lam & Fresco, 2015).

Current methods to identify and understand non-adherence have major limitations. Direct observation may be intrusive or expensive, and indirect observation through patient surveys relies heavily on a patient’s capacity to remember and report adherence to medication treatment. Our ultimate goal is to study for reasons for non-adherence using social media data at a large scale, as it is generally inexpensive, unintrusive, and does not rely on a patient’s memory of events in the distant past. To the best of our knowledge, the few studies on non-adherence using social media restrict their search to health forums dedicated to long-term or chronic conditions. This choice helps to process and interpret the data but greatly reduces the size of their corpora and, therefore, limits the types of reasons discovered. With 321 million active users per month in 2019 (Shaban, 2019), Twitter can prove to be a valuable source of data for non-adherence studies that scale.

In preliminary work (Onishi, et al., 2018), we found that, given the micro-blogging format of Twitter, many users report changes to their medication treatment in tweets separate from those reporting the reasons for those changes, i.e., that the mention of a change in treatment does not always provide evidence of non-adherence. Thus, we approach the detection of non-adherence on Twitter in two stages: (1) the tweet-level detection of changes to medication treatment, and (2) the user-level analysis to determine whether the reasons for change could be understood as non-adherence. This study focuses on the first stage, aiming to automatically detect tweets in which users report changes to their treatment, regardless of whether the changes were recommended. Automatically detecting tweet-level reports of changes to medication treatment enables the large-scale use of Twitter data for studying non-adherence at the patient level.

The main contributions of our work are (1) the release of two corpora collected from social media, manually annotated with medication change, (2) a binary classifier based on neural networks to detect changes in treatment, and (3) a manual analysis of non-adherence reasons expressed in Twitter for a general set of drugs.

## 2. Related Work

Prior work has focused on attempting to find non-adherence mentions and reasons for non-adherence from different data sources. For this purpose, researchers have mined the unstructured portion of clinical notes (Sohn, et al., 2010; Topaz, et al., 2016) or messages from clinical portals (Yin, et al., 2018). However, these documents are protected, and they do not routinely contain self-reported non-adherence written by patients. Automatically extracting reasons for non-adherence is harder from social media data. An intuitive approach to tackle this difficulty is to manually analyze a sample of posts and this has been done with data from health forums, Facebook, and Twitter (Mao, et al., 2013; Bhattacharya, et al., 2017). However, larger studies can benefit from Natural Language Processing methods, to at least automatically filter relevant from irrelevant posts and reduce annotation burden, and to analyze large amounts of data. Unsupervised methods are attractive for the latter, since they require few or no annotations to learn the task, relying on topic modeling (Abdellaoui, et al., 2018) or interactive exploration of the data with search engines (Bigeard, et al., 2019). In general, unsupervised approaches resulted in high recall but low precision. Despite the challenges of annotation (Belz, et al., 2019), supervised methods gave the best results. Bigeard et al. (Bigeard, et al., 2019) achieved 0.824 F1-score on health forums data in French with a Naïve Bayes and hand-crafted features approach. Comparable performance was observed also on health forums, 0.882 F1-score, reported by Yin et al. (Yin, et al., 2017) with a binary logistic regression model working with word embeddings-based features. Both studies used 1,000 or fewer annotated examples. In (Xie, et al., 2017), the authors collected drug reviews written by users of the health forum WebMD^1^. They applied a binary classifier to detect sentences mentioning non-adherence, then a sequence labeler to extract the reasons in 4,500 reviews. The performance of their classifier, a bidirectional Long Short-Term Memory neural network, when trained on 8,000 examples and tested on 2,000 examples was 0.828 F1-score. These performances are, however, aided by the fact that they were working on health forums where users focus their discussions on medical issues. In this context, phrases are unambiguous and automatic systems can learn reliable linguistic patterns. A further limitation of these studies is that it is unclear whether they validated if the stated change in medication regimen was done with or without a doctor’s approval. This is a defining characteristic of non-adherence that is particularly challenging to establish, even for human annotators. Thus, in this study, our main interest is to detect medication treatment changes in Twitter. This is much less ambiguous and enables the deployment of automatic methods on a much larger scale. The volume of reports that can be collected on Twitter could provide a broader view of non-adherence behaviors (e.g. stopping a treatment because of an adverse drug event experienced or feared). Moreover, in Twitter, non-adherent users could potentially be directly contacted and invited to participate in a study. In contrast, following up with the users would not be possible in WebMD since they post reviews anonymously. However, compared to forums, Twitter poses a new challenge for automatic detection. Whereas 55% of reviews mention a change in our WebMD corpus (see Section 3.1.1), in a separate study (Golder, et al., 2020) we sought to ascertain the topics discussed by users of statins, annotating a corpus of 12,649 tweets that mention a statin, and found that only 1.9% of them (251 tweets) mentioned a change in medication treatment. Such sparsity of positive examples makes the collection of training examples difficult and degrades the performance of learning algorithms (Weissenbacher, et al., 2019). We addressed this problem by training our classifiers with a common training strategy: under-sampling the negative examples (Haixiang, et al., 2017).

## 3. Methods

Our first effort was to collect corpora suitable to train our classifiers with supervision. Knowing that mentions of medication changes in Twitter are rare and that training corpora for supervised methods usually contain several thousand positive examples, we judged the cost too high to create a balanced training corpus. We opted for an alternative solution. We took advantage of two training approaches to reduce the annotation effort, transfer and active learning (Kasai, et al., 2019). We explain these approaches in detail in this section.

### 3.1 Corpora

We detail two corpora collected for this study in this section and summarize their statistics in Table 1.

**Table 1:** Corpora Statistics.

| COUNTS PER CORPUS |  |  |
| --- | --- | --- |
|  | WebMD | Twitter |
| TRAINING SET | 10,378 reviews (5746 + / 4632 -) | 5,898 tweets (518 + / 5380 -) |
| VALIDATION SET | 1,297 reviews ( 741 + / 556 -) | 1,572 tweets (138 + / 1434 -) |
| TEST SET | 1,297 reviews ( 735 + / 562 -) | 2,360 tweets (208 + / 2152 -) |
| **TOTAL** | **12,972 reviews** | **9,830 tweets** |

#### Twitter Corpus

We collected 9,830 tweets to train and evaluate the classifiers used for this study. We combined tweets mentioning drugs from two existing corpora: (1) the corpus released during the first track of the SMM4H’18 shared-task (Weissenbacher, et al., 2018), a Natural Language Processing competition to identify drug mentions, and (2) a subset of 1,367 tweets, randomly selected from the corpus collected by Golder et al. (Golder, et al., 2020). Two annotators labeled all tweets with “1” if the tweets mention a change in the medication treatment, “0” otherwise. Based on double-annotating 4,931 of the 9,830 tweets, their inter-annotator agreement was 0.65 Cohen’s Kappa score. This inter-annotator agreement score reflects a moderate agreement between the annotators (McHugh, 2012), implying that they had to rely often on their common and medical knowledge to label the tweets. Annotators followed our guidelines, available at XXX.

#### WebMD Corpus

Our second corpus consists of reviews written by anonymous users on WebMD. This website provides an opportunity for users to review drugs. A review is assigned to exactly one drug and is composed of three scores evaluating the satisfaction, effectiveness, and ease of use of the drug. The scores range from 1 to 5 stars, with 1 star being the lowest value. Users can also comment on their personal experience with the drug in a free text form. The comment is optional and limited to 2,000 characters. In August 2018, we collected all reviews from WebMD using in-house software. We randomly sampled 12,972 reviews with a comment and 1 or 2 stars for satisfaction. We selected posts with low satisfaction scores since unsatisfied users are more likely to stop or to change medication. As for the tweets, two annotators labeled the reviews with “1” if a review mentions a change of medication, “0” otherwise. The inter annotator agreement was also moderate with 0.74 Cohens’ Kappa score (McHugh, 2012).

### 3.2 Classification Approaches

#### 3.2.1 Baseline

We implemented a simple rule-based classifier as a baseline. It applies a set of regular expressions on a corpus and every post matched by one expression was labeled as mentioning a medication change. We designed two sets of regular expressions manually, one on the training set of the WebMD Corpus and one on the training set of the Twitter Corpus. We evaluated this classifier on both test sets of the corpora.

#### 3.2.2 Convolutional Neural Network

We selected Convolutional Neural Networks (CNNs) to detect posts stating a medication change. We ran experiments involving active learning which requires training several networks during multiple iterations (Lu, et al., 2019). In this setting, CNNs present an advantage over more complex networks as they are fast to train.

Our CNNs use a standard architecture to compute the probability for an input post to mention a medication change. A CNN accepts a 400×100 matrix as input, where 100 is the number of words of a post to classify and 400 is the dimension of the embeddings representing each word of the post; 100 is a fixed length, right padded for shorter posts, and right truncated for longer ones. We chose existing word embeddings, pre-trained on 400 million tweets with word2vec (Godin, et al., 2015). We assigned vectors randomly initialized for out-of-vocabulary tokens, fine-tuned during training. We used dropout on the input and hidden layers of the CNN to avoid overfitting. We used a RELU activation function for all appropriate layers, except for the last layer, where we used a Softmax function. We implemented only one convolutional layer in our CNN with 400 filters, a kernel of size 3, and stride 1. We preprocessed our posts with a TweetTokenizer from the Natural Language Toolkit (Bird, et al., 2009). We removed username handles from the tweets, reduced elongated words, and lowercased all posts. Our code is publicly available^2^ and details all hyperparameters.

#### 3.2.3 Convolutional Neural Network with active and transfer learning

As a way to improve the training of our CNNs despite the strong imbalance between the tweets mentioning and not mentioning medication change in the Twitter Corpus, we used active and transfer learning. Our experiments involved a large number of hyperparameters chosen based on our prior experience. Their optimization through grid-searches is left as future work.

##### Active learning

denotes a family of supervised learning algorithms (Settles, 2012) for training a classifier with a limited number of annotated examples. In a standard supervised learning algorithm, all examples annotated available are used to train a classifier. In active learning, an artificial agent, the learner, is introduced to select, from a pool of unlabeled examples, which examples should be annotated for training a classifier – or an ensemble of classifiers. Whereas a passive learner randomly selects the examples, an active learner has an algorithm to select the most relevant examples. Intuitively, the active learner focuses on unlabeled examples likely to be incorrectly classified by its classifier (see Figure 1). Such examples may be located close to the decision boundary of the classifier, revealing the need to update the parameters of the classifier’s model (e.g. changing the slope of a linear model) or they may be located in new areas in the features space, revealing the need to change the classifier’s model itself (e.g. changing to a non-linear decision boundary). An active learner learns a task with fewer examples than a passive learner since it queries only for examples resulting in useful changes of the classifier’s model and ignores redundant examples.

**Figure 1:**
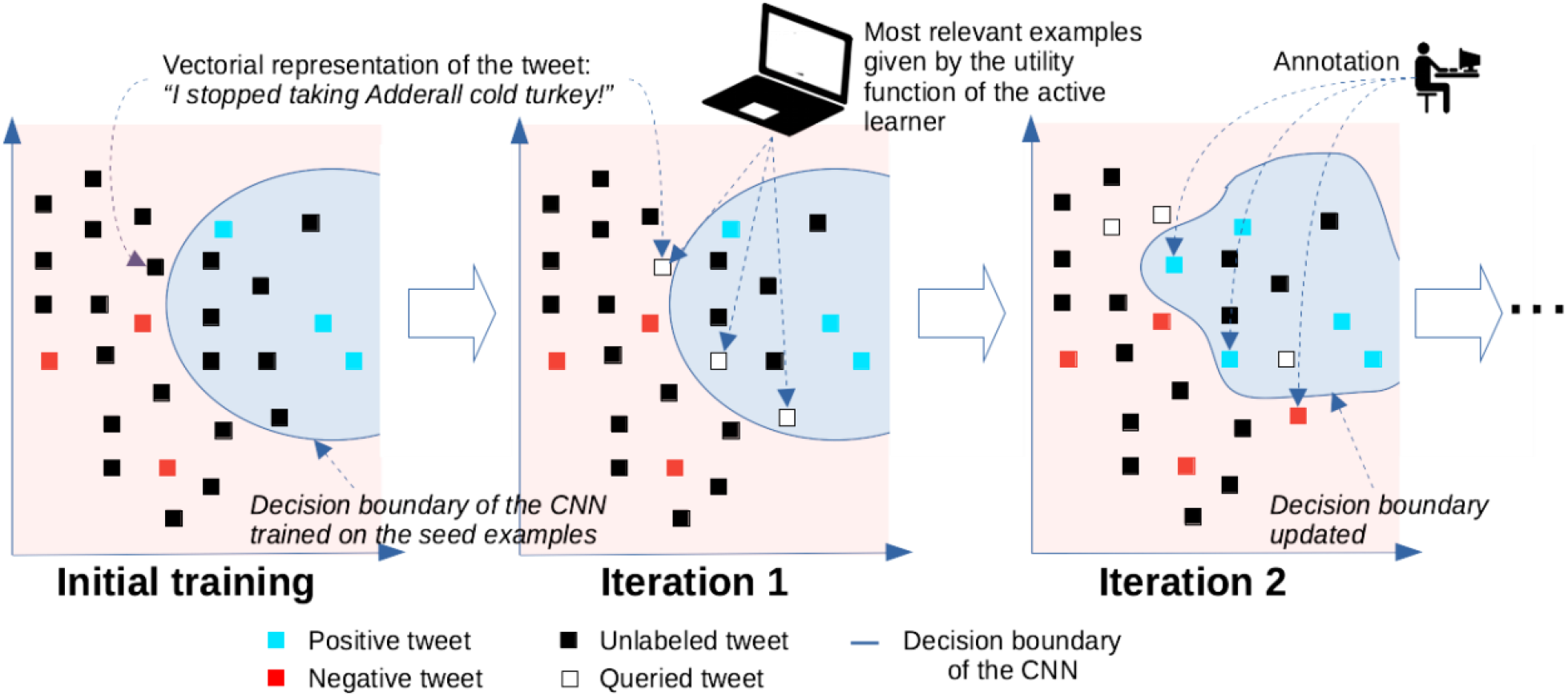
Two iterations of the active learning algorithm. All Tweets in the blue zones delimited by the decision boundaries are labeled as positive by a CNN classifier, tweets in the red zone are labeled as negative. The decision boundaries defined by the models of the classifier are updated at each iteration to include newly labeled positive tweets. (Note: we reduced the dimensions of the vectors to two dimensions for representation purpose)

We first evaluated the benefits of active learning alone, on the WebMD corpus and on the Twitter corpus. We compared three standard utility functions to select the most relevant examples: random sampling as a baseline, uncertainty sampling, and disagreement sampling with a committee of 5 CNNs randomly initialized. For uncertainty sampling and disagreement sampling, we chose the entropy and the vote entropy to measure the uncertainty of the learner (Settles, 2012). Our setting was identical for all experiments. We kept 10% of the training set as seed set and placed the remaining examples in a pool, with their labels hidden from the learner. The steps of the active learning algorithm are as follows:

a. Our learner started its initial training on the seed set.
b. The learner queried the labels for 200 new examples from the pool using its utility function. All examples of our corpus being pre-labeled, we simply released their labels to the learner.
c. The learner updated the model of its classifier with the 200 new examples and evaluated its performance on the validation set as well as on the test set and removed these 200 examples from the pool.
d. The learner iterated the steps (b), (c), and (d) until no example remains in the pool.

Figure 1 shows the step (a) and two iterations of steps (b), (c), and (d). The learner saved its classifier’s models at each iteration (c) and selected the model *m* at the iteration where it achieved its best performance on the validation set. We evaluated the learner on the test set using the model *m*. The model *m* may not be the model which obtained the best performance on the test set, but choosing *m* is an effective heuristic to find a model appropriate for real applications.

##### Transfer learning

is a heuristic to improve the performance of a supervised classifier (Kasai, et al., 2019). With transfer learning, a classifier solves a new task by reusing in its inference the knowledge it acquired when solving a similar task. In the case of a neural network classifier, the knowledge is instantiated by the weights of the network.

We evaluated the benefits of transfer learning by implementing a passive learner using a single CNN. We started by training our classifier on all training examples of the WebMD corpus - the *source corpus* - to learn the linguistic patterns indicating a change in the medication treatment. We transferred its knowledge by continuing the training of the classifier with all training examples from the Twitter corpus - the *target corpus*. We evaluated our classifier on the test set of the Twitter corpus. We also measured the loss when our classifier did not have past knowledge. The CNN was only trained and evaluated on the target corpus. Considering the similarity between the source and the target corpora, we checked the performance of a CNN first trained on the training source corpus and, with no additional training step on the training set of the target corpus, evaluated on the test set of the target corpus.

##### Transfer and Active Learning

were used in combination and evaluated with the same settings as for evaluating transfer learning, with one difference, we used an active learner during the training phases of the classifier on the source and/or target corpora.

All classifiers and learners evaluated during our experiments are summarized in Table 2.

**Table 2:** Classifiers and learners summary.

| System | Description | Experiment |
| --- | --- | --- |
| RegEx | Classifier using hand-crafted regular expressions | RE1 – Expressions crafted on WebMD training set; evaluated on WebMD test set<br>RE2 – Expressions crafted on Twitter training set; evaluated on Twitter test set |
| Random | Passive Learner using a single CNN and random sampling | RA1 – Trained on WebMD training set; evaluated on WebMD test set<br>RA2 – Trained on Twitter training set; evaluated on Twitter test set<br>RA3 – Trained on WebMD training set, with/without update on Twitter training set; evaluated Twitter test set |
| Uncertainty | Active Learner using a single CNN and uncertainty sampling | UN1 – Trained on WebMD training set; evaluated on WebMD test set<br>UN2 – Trained on Twitter training set; evaluated on Twitter test set<br>UN3 – Trained on WebMD training set, with/without update on Twitter training set; evaluated Twitter test set |
| Committee | Active Learner using a committee of 5 CNNs and disagreement sampling | CO1 – Trained on WebMD training set; evaluated on WebMD test set<br>CO2 – Trained on Twitter training set; evaluated on Twitter test set<br>CO3 – Trained on WebMD training set, with/without update on Twitter training set; evaluated Twitter test set |

### 3.3 Validation

Using our classifier, we collected a large corpus of tweets mentioning a change of medication treatment and manually analyzed the tweets to determine whether users were non-adherent to their prescriptions and the reason why, if given.

We collected 1,936,820 tweets from January 2019 to April 2020. We queried the stream of Twitter using the official application programming interface to retrieve tweets mentioning drug names, or their variants, from a pre-defined list of 1,322 drugs. The 1,322 drugs were randomly selected from the RxNorm database (https://www.nlm.nih.gov/research/umls/rxnorm/overview.html, accessed June 11, 2018). We applied our best classifier (see Section 4.1.2) on these 1.9 million tweets and detected 5,811 tweets with a probability equal or higher than 0.95 to mention a change of medication. From this 5,811, we manually analyzed a subset of 3,010 tweets, randomly selected. One annotator first confirmed the decision of our classifier - the tweet mentioned a medication treatment change - and then looked for the reasons of this change in the timeline of the user, up to ten tweets posted before and after the tweet mentioning the change as previously done in (Onishi, et al., 2018). If the tweet was a part of a discussion, the annotator also looked for the reasons into the discussion thread. We determined non-adherence if it was stated, or could be inferred, from the tweet that the user changed or stopped taking the medication without consulting their provider. For example, in the tweet, “*took Prozac for a while, took myself off it didn’t like the side effects from it*”, the user is clearly stating that they made the decision (‘took myself off it’) rather than their doctor, thus it was labeled as non-adherence. Our senior annotator (K.O) categorized the reasons for non-adherence and our experts in pharmacoepidemiology (SH and RG) validated a subset of them.

## 4. Results

### 4.1 Automatic detection of medication change

We detail our results in Table 3. We trained and evaluated our classifiers/learners five times and reported the means of their precision, recall, and F1 scores to reduce the differences caused by their stochastic optimizations. True Positives (TP) are posts that mention a change in treatment and are detected by a classifier. False Negatives (FN) are posts that mention a change in treatment but are not detected by a classifier. False Positives (FP) are posts that do not mention a change in treatment but are detected by a classifier. The Precision is the fraction of posts correctly classified as positive among all posts classified as positive: TP/(TP+FP). The Recall is the fraction of posts mentioning a change successfully retrieved: TP/(TP+FN). The F1 score is the harmonic mean of the precision and recall; it summarizes the overall performance of the classifier.

**Table 3:** Binary classification of medication change in WebMD and Twitter posts. (Precision, Recall and F1 scores are given in percentage; classifiers trained by an active learner are marked with italic fonts)

| Corpus: WebMD |  |  |  |  |
| --- | --- | --- | --- | --- |
| System |  | Precision | Recall | F1 |
| RE1 – RegEx |  | 68.3 | **90.3** | 77.8 |
| RA1 – Random |  | 78.7 | 85.9 | 82.1 |
| *UN1 – Uncertainty* |  | **81.4** | 83.1 | 82.2 |
| *CO1 – Committee* |  | 80.2 | 85.7 | **82.8** |
| Corpus: Twitter |  |  |  |  |
| System |  | Precision | Recall | F1 |
| No Transfer | RE2 – RegEx | 45.3 | 41.4 | 43.2 |
|  | RA2 – Random | 50.8 | 34.1 | 40.4 |
|  | *UN2 – Uncertainty* | 39.3 | **56.2** | 45.5 |
|  | *CO2 – Committee* | 51.7 | 34.7 | 41.3 |
| Transfer with and without update on target corpus | RA3 – Random | 53.6 / 38.7 | 40.9 / 45.5 | 46.2 / 41.3 |
|  | *UN3 – Uncertainty* | 46.5 / 26.4 | 52.4 / 51.6 | 48.4 / 34.9 |
|  | *CO3 – Committee* | **56.2** / 30.7 | 45.8 / 52.5 | **50.4** / 38.7 |

The drop of performance for all classifiers when applied on our Twitter corpus stands out in Table 3. Whereas the learner with a committee of CNNs, the best learner on both corpora, achieved a score close to human performance on the WebMD corpus, with 0.83 F1-score, its performance dropped from 0.83 to 0.50 F1-score on the Twitter corpus. This drop is likely due to the difference of genre between our two corpora (Poibeau, 2003). When users are reviewing drugs in WebMD, they only discuss their experiences and rarely diverge. We can express few generic and reliable patterns on this corpus to detect medication change, e.g. the phrase *“side effect”* most often indicates a change. When users are posting on Twitter, they discuss other subjects than their medication experience. Such generic patterns become unreliable. To improve their precision, we needed to integrate more constraints to model the surrounding context in tweets, and consequently, to keep a high recall, multiplying close variation of the patterns. Such adaptation of the patterns remains challenging for both humans and classifiers (Vanni, et al., 2018).

#### 4.1.1. Reducing annotation effort with Active Learning

We report in Figure 2 the average performances on the test sets of our two corpora of our three learners when trained on various subsets of the training examples. Active learning helped to accurately perform our task faster, reducing the need for additional training examples. The active learners reached their best performances plateau earlier than the baseline passive learner on both corpora. Active learning did not cause a significant drop in performance compared to learning on all examples available. We marked with black circles the average percentage of the training examples the learners analyzed when they achieved their best performance on the evaluation sets during the five runs. The models selected, achieving the best performance on the validation sets, achieved lower but close performances from the best models on the test sets and were in their vicinity. It is difficult to explain this result, but this chart illustrates a general tendency of our active learners; they all tend to achieve their best performance earlier on the validation set.

**Figure 2:**
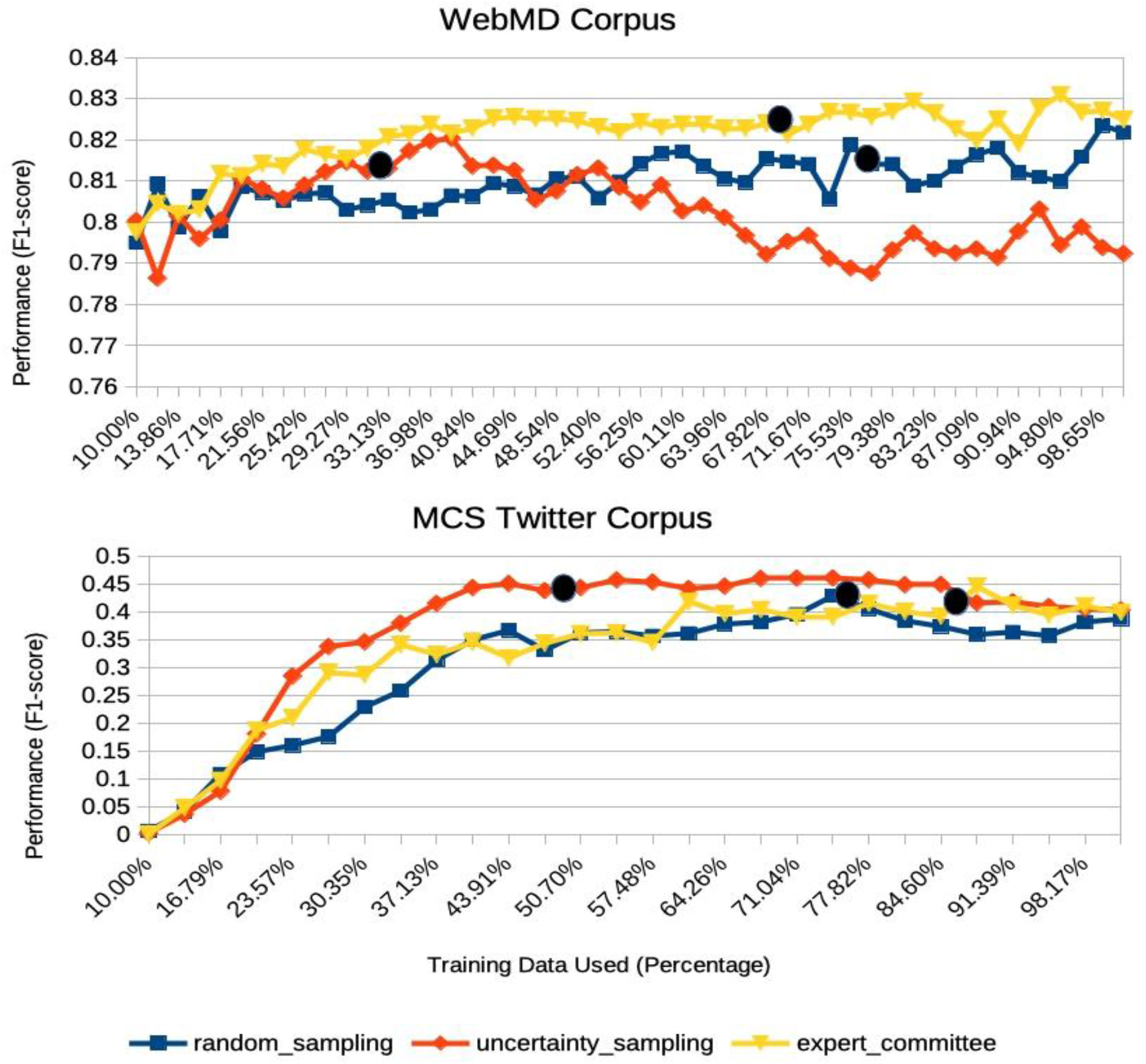
Classifiers’ performance on WebMD and Twitter corpora test sets with increasing training data. (Black circles indicate the average percentage of training examples analyzed by the classifiers when achieving their best performance on validation sets)

#### 4.1.2 Benefits of transfer learning

We achieved our best score on the Twitter corpus, 0.50 F1-score, with an active learner using a committee of CNNs and transfer learning. Because users express their change of medication with the same linguistic patterns in social media, our learner could learn the patterns from the WebMD corpus, a larger and more balanced corpus, and performed more efficiently the task on the Twitter corpus. The transfer allowed an increase of 9.1 F1-score points for the committee. A requirement, however, is to adjust the reliability of each pattern on the target corpus through additional training iterations on a small sample of examples from the target corpus. Without adapting its classifiers’ models, there was no evidence our learner gained anything from the transfer as shown by the experience CO3 in Table 3.

### 4.2 Analysis of reasons for medication non-adherence

Our manual analysis of the subset of 3,010 tweets, detected by our best classifier as very likely to mention a medication change, confirmed our preliminary results published in (Onishi, et al., 2018): users post about their medication non-adherence on Twitter and were likely to explain their reasons in the tweets assessed (including previous and subsequent tweets). From these 3,010 tweets, 1,956 were True Positive (i.e., tweets mentioning a medication change). Among these 1,956 tweets, 19.2% (375) were explicitly mentioning non-adherence with the reasons explained in the tweets themselves (68%, 255) or their contexts (9.1%, 34). In this example, *“i was taking my adderall less than prescribed to save money, […]”*, the non-adherence reason was categorized as Access Issue due to cost. Out of 375, only 86 tweets (22.9%) did not have a reason indicated in their context.

Table 4 summarizes the classes of stated reasons for non-adherence. Adverse drug reaction was the most common reason mentioned for being nonadherent, with 19.2% (72) of our tweets. Misuse and abuse were the second reason with 17.8% (67). This reason could be over-represented in our sample because 57.9% (217) of the tweets mentioning non-adherence are mentioning mixed amphetamine salts Adderall, a drug abused for its stimulant properties. Access issues to the drugs, caused for example, by their costs or problems with insurances and refill issues, were also a major concern to patients and counted for the third reason of non-adherence with 12.8% (48), before unintentional non-adherence of users, only 6.4% (24).

**Table 4:** Reasons of non-adherence discovered in a sample of tweets mentioning drugs. Statements are unedited for spelling, punctuation, or format.

| NON-ADHERENCE REASON | DESCRIPTION / EXAMPLE | COUNT (PERCENT) |
| --- | --- | --- |
| **ADVERSE DRUG REACTION** | Experienced/fear of adverse drug reaction<br>*"I hate avapro after a few doses I got a sore throat that lead to nonstop coughing. I stopped taking it two days ago. [...]"* | 19.2 (72) |
| **MISUSE/ABUSE** | Indication that the medication was being abused/misused<br>*"[...] when I abused adderall now and then for a while [...]"* | 17.8 (67) |
| **ACCESS ISSUES** | Unable to get medication (cost, insurance, refill issues etc.)<br>*"I stopped taking my Lamotrigine and took myself off quetiapine bc I no longer wanted to pay for them [...]"* | 12.8 (48) |
| **BELIEFS** | Various beliefs (not needed, being overmedicated, harmful etc.)<br>*"Yes big pharma is the reason I stopped taking my Xanax and other meds"* | 11.2 (42) |
| **UNINTENTIONAL** | Non-adherence seems unintentional (forgotten, error dosage etc.)<br>*"I didn't take my Adderall for a while because I lost my bottle and today was my first day back on it a [...]"* | 6.4 (24) |
| **PARENT'S DECISION** | Parent took child off medication<br>*"my dad's taking me off of my vyvanse prescription without my doctors advise and self-undiagnosing me even though he only took me off meds bc of my side effects from letting me have weekends off"* | 2.4 (9) |
| **EFFICACY** | Not effective or higher dosage needed<br>*"When the pain med stopped working, I stopped going They gave me no other treatments. Was wasting my time!"* | 2.3 (8) |
| **STIGMA** | Patient felt a stigma being on medication<br>*"The doctor treated me like a drug addict, [...] I discontinued the med because of the shame"* | 1.3 (5) |
| **OTHER** | Did not fit into any of above categories<br>*"[...] He has been neglecting caring for himself since finding out he couldn't get on the transplant list. He stopped taking his diuretic. [...]"* | 3.7 (14) |
| **NOT AVAILABLE** | Reason not found in tweet context<br>*"I stopped taking ramipril on my own terms weeks ago."* | 22.9 (86) |
| **TOTAL** |  | 100 (375) |

## 5. Discussion

In this study, our objective was to automatically detect tweets mentioning changes in medication treatment, and manually analyze their context to assess whether the reasons of the changes were given and whether we could determine if they were advised by a physician or not.

Our classifier for detecting medication changes in WebMD reviews obtained an F1-score of 0.828, a score equal to the score of Xie’s classifier (Xie, et al., 2017). However, we cannot compare directly the two scores since we are classifying medication treatment changes in general and not directly non-adherence. In addition, Xie’s corpus includes WebMD reviews with positive and negative scores. Positive reviews are less likely to mention medication changes and are thus easier to classify. Including positive reviews may have artificially inflated their score. Their dataset was not made available, so this is impossible to verify. Our intent for using WebMD was to transfer the linguistic knowledge easily acquired on this balanced corpus.

Transfer and active learning appear to be efficient heuristics to help train classifiers on extremely imbalanced corpora. Compared to a traditional supervised learning approach, we were able to reduce the number of labeled training examples while increasing the performance of our learner by 9.1 points in F1-score by combining both heuristics. These positive results were obtained on one task and need to be confirmed. We intend to repeat our experiments on the adverse drug event detection and drug detection tasks described in (Weissenbacher, et al., 2019; Weissenbacher, et al., 2019).

We made two experimental choices to facilitate our evaluation of transfer and active learning. These choices limited our performance. We used a well-established neural network architecture for our classifiers and well-known utility functions for our active learners, whereas better alternatives already exist. Such an alternative could be a dense representation of the entire posts using ELMo or BERT neural networks (Peng, et al., 2019). With this representation, it also becomes possible to express new utility functions. When neural networks encode the posts, they encode their semantic and place similar posts close to each other in a multidimensional space and posts with different meanings far away. Such representation could help an active learner to explore unlabeled data by clustering posts expressing the same meaning despite lexical and syntactical variations (Lu, et al., 2019; Kholghi, et al., 2016).

In this study, we hypothesized that the linguistic patterns expressing medication change in social media are similar across different drug classes. We focused our efforts on developing a general learner by training it on corpora composed of posts mentioning any medication. To our knowledge, no prior work exists that targets medication change mentions in social media. However, in general, epidemiological studies in pharmacovigilance include only a particular class of drugs (rather than a random collection of them) to discover unknown reasons for non-adherence. The transfer learning approach presented in this study can be used on a new corpus of tweets mentioning a specific drug class of interest by simply retraining our classifier on this new corpus as it was done in this study for the general collection of drugs. Future work in this direction will benchmark the performance of the classifier per medication class (for example, for statins or anti-hypertensives).

## 6. Conclusion

In this study, we presented an ensemble of CNNs to detect tweets mentioning changes in the medication treatment. Using transfer and active learning, we achieved 0.50 F1-score, a score high enough to collect a large number of tweets of interest and manually analyze their context to determine if users were nonadherent to their prescriptions. We conclude that Twitter users do state their non-adherence to a medication treatment and are likely to explain their reasons in their timelines, suggesting that Twitter data, systematically collected and automatically analyzed, could supplement current efforts in identifying patient-stated reasons for non-adherence. A major challenge remains to fully automate the detection of non-adherence and their reasons for larger studies.

## Data Availability

All data will be released during the text mining competition #SMM4H'21 at NAACL'21

https://healthlanguageprocessing.org/smm4h/

https://2021.naacl.org/

## Footnotes

1 Available at: https://www.webmd.com/drugs/2/index

2 https://bitbucket.org/davy-weissenbacher/medchange/src/master

